# Discover overlooked complications after preeclampsia from three real-world medical record datasets of over 100,000 pregnancies

**DOI:** 10.1101/2023.12.05.23299296

**Authors:** Haoming Zhu, Xiaotong Yang, Leyang Tao, Wanling Xie, Jui-Hsuan Chang, Zhiping Paul Wang, Elizabeth Langen, Ruowang Li, Lana X Gamire

## Abstract

**Importance:** Preeclampsia poses a significant threat to women’s long-term health. However, what diseases are affected and at what level they are affected by PE needs a thorough investigation.

**Objective:** To conduct the first large-scale, non-hypothesis-driven study using EHR data from multiple medical centers to comprehensively explore adverse health outcomes after preeclampsia

**Design:** Retrospective multi-cohort case-control study

**Participants:** We analyzed 3,592 preeclampsia patients and 23,040 non-preeclampsia controls from the University of Michigan Healthcare System. We externally validated the findings using UK Biobank data (443 cases, 14,870 controls) and Cedar Sinai data(2755 cases, 60,305 controls).

**Main outcomes:** We showed that six complications are significantly affected by PE. We demonstrate the effect of race as well as preeclampsia severity on these complications.

**Results:** PE significantly increases the risk of later hypertension, uncomplicated and complicated diabetes, renal failure and obesity, after careful confounder adjustment. We also identified that hypothyroidism risks are significantly reduced in PE patients, particularly among African Americans. Severe PE affects hypertension, renal failure, uncomplicated diabetes and obesity more than mild PE, as expected. Caucasians are affected more negatively than African Americans by PE on future hypertension, uncomplicated and complicated diabetes and obesity.

**Conclusion:** This study fills a gap in the comprehensive assessment of preeclampsia’s long-term effects using large-scale EHR data and rigorous statistical methods. Our findings emphasize the need for extended monitoring and tailored interventions for women with a history of preeclampsia, by considering pre-existing conditions, preeclampsia severity, and racial differences.

**Key Points:** *Question:* Which complications are more or less likely to occur in patients after preeclampsia (PE), compared to those with normal pregnancies? How do the severity of PE and patient race further affect these complications?

*Findings:* In a retrospective case-control study on 3 datasets (n = 26,632, n = 15,313, n= 63,060), we identified 5 complications with significantly increased risk due to PE, including hypertension, uncomplicated and complicated diabetes, renal failure and obesity, after careful confounder adjustment. We also identified that hypothyroidism risks are reduced in PE patients, particularly significant among African Americans. Severe PE affects hypertension, renal failure, uncomplicated diabetes and obesity more than mild PE, as expected. Caucasians are affected more negatively than African Americans by PE on future hypertension, uncomplicated and complicated diabetes and obesity.

*Meaning:* Patients with a history of PE should be closely monitored and carefully assessed for the risks of developing these complications.

## Introduction

Preeclampsia (PE) is a severe pregnancy complication that emerges after the 20th week of gestation and is characterized by hypertension and end-organ damage^1^. Its prevalence in the United States is estimated at approximately 4%, with a global prevalence of 2% to 8%^2,3^. PE profoundly impacts maternal and neonatal health, primarily through a hypertensive state that increases vascular resistance, potentially impairing blood flow to essential organs^4^. This condition leads to endothelial dysfunction, which can cause extensive damage to the mother’s kidneys, liver, and central nervous system^5^. Additionally, PE adversely impacts placental perfusion, potentially resulting in fetal growth restrictions and preterm births^6^. While the resolution of pregnancy often mitigates the acute symptoms of PE, its long-term-lasting effects on maternal health, manifesting as various chronic complications such as hypertension, cerebrovascular disease, diabetes, cardiovascular disease, and renal disease, can persist for years postpartum^7^.

Despite the efforts to study the long-term effect of PE with hypothesis-based case-control studies, large-scale non-hypothesis-based comprehensive discovery of complications after PE with rigorous statistical analysis is currently lacking. Previous cohort and meta-analysis studies were mostly hypothesis-based, focusing on confirming known associations and may overlook important associations^8–13^. Additionally, most studies either did not adjust for confounders or only accounted for basic demographics, leaving out the important confounders like pre-existing health conditions^8–16^. Moreover, few studies have paid attention to the severity and racial disparities in PE’s long-term effects. As statistical methods evolve, a more robust and comprehensive approach utilizing modern analytical methods and large datasets is imperative to explore the long-term effects of PE and offer clinical insights.

With the widespread use of electronic medical systems, the Electronic Health Record (EHR) has become an important source of patient data for medical research. EHR provides comprehensive and timestamped patient information in large volumes, including demographics, diagnosis records, social history, laboratory results, etc. These features make it compatible with modeling quantitative outcomes, such as patient survival. Survival analysis enables the examination of time until the occurrence of specific events^17^. Compared with basic statistical models such as t-tests and linear regression, survival analysis offers advantages in terms of handling censorship, accommodating non-normally distributed data, and providing a more detailed understanding of disease trajectories. Incorporating survival models into the analysis offers a better understanding of PE’s long-term consequences.

This study adopts a non-hypothesis-based exploration, leveraging EHR data to investigate connections between PE and subsequent health trajectories. Our goals are: (1) to identify and externally validate any significant complications after PE; (2) to comprehensively study their trajectories by severity of PE, and (3) to reveal potential racial disparities. We aim to enhance the clinical understanding of PE’s long-term consequences and ultimately help care providers adopt preventive aftercare strategies.

## Methods

### Data Source

We fit and externally validated the models using EHR data from three large databases. The discovery data are from the *University of Michigan (UMich) Medicine Healthcare System*, an academic health system in Michigan State. The University of Michigan Medical School’s Institutional Review Board (IRB) granted data utilization approval under HUM#00168171. The EHR from the *UM Medicine Healthcare System* provides comprehensive features, including diagnoses, encounter information, demographics, medications, etc. The complete records for these features are available starting in 2006^18^. We obtained the first validation data from the *UK Biobank*, a long-term study with approximately 500,000 volunteers, providing similar features^19^. All *UK Biobank data* for the confirmation cohort pertained to project 86494. These records are available starting in 2006. The second validation dataset comes from Cedar Sinai Hospital. The Cedar Sinai limited PHI database includes EHR data with similar features collected from 4.9 million patients since 2011. Our study initially utilized features including diseases diagnosed, age at diagnosis, pre-existing medical conditions, race, and social history.

The discovery data includes case patients with at least one PE diagnosis between 2003 and 2023 and controls with at least one pregnancy during the same period and no PE or other PE-related diseases (PE, eclampsia, pre-existing hypertension complicating pregnancy, gestational edema, or maternal hypertension). Diagnoses were based on the International Classification of Diseases (ICD)-9 and -10 codes^20,21^ (**Supplementary Table 1**). We excluded patients with no social history or pre-pregnancy medical history to avoid introducing biases in the statistical modeling. Patients with missing or unknown race information are coded as “others” in race. We followed the same selection criteria to select the *UK Biobank* cohort *and Cedar Sinai* cohort.

### EHR feature engineering

As a non-hypothesis-based study, we considered all diagnosis records within 10 years after delivery as potential targets. We exclude temporary postpartum conditions within 90 days of delivery to focus better on long-term health outcomes. We then classified these records into 30 medical complications, such as uncomplicated hypertension, complicated diabetes, obesity, etc., according to the Elixhauser Comorbidity Index^22^. The Elixhauser comorbidity index is a widely used method in healthcare to comprehensively assess and quantify the severity of multiple health conditions a patient might have^23^. Although this index is not disease-specific, it significantly improved the feasibility of non-hypothesis-based analysis by reducing numerous diagnosis codes into 30 categories. A complete list of matches between ICD-10 codes and Elixhauser Comorbidities is shown in **Supplementary Table 2**.

To ensure the rigor of our analysis, we adjusted for various confounders including patient age, race, smoking status, alcohol usage, and pre-pregnancy health conditions. Pre-pregnancy complications are defined as diseases presented at least 200 days before their first PE (for cases) or their first normal pregnancy (for controls) record. Adjusting for pre-pregnancy complications is pivotal to avoid conditions related to or comorbid by preeclampsia. We encoded the pre-pregnancy complications as binary entries (1 for presence, and 0 for absence) following the Elixhauser Comorbidity Index. We removed features with more than 20% missing values or a p-value > 0.10 in the initial univariable test as advised^24^.

### Non-hypothesis-based PE complications identification

We first identified the significant complications after PE out of all 30 Elixhauser comorbidities by calculating the cumulative risk of each disease at intervals of one year, two years, and up to ten years following PE or normal pregnancy. The follow-up length for each patient was defined as the time between their most recent record and their first PE diagnosis (for cases) or normal pregnancy (for controls). For each disease, we calculated odds ratios (OR) at N years post-PE using logistic regression with adjustment of aforelisted confounders^25^. OR is a measure of the association between the presence of a particular condition and a specific outcome, expressing the odds of the event occurring in the cases relative to those in the controls^26^. We considered the complications that show significance in OR (p < 0.05) in more than 4 consecutive years to be significantly associated with PE. We calculated the effective size and Variance Inflation Factors (VIF) of each covariate using the package “qacReg” to exclude covariates with low effective size and potential collinearity among independent variables. The final features of each model are listed in **Supplementary Table 3**.

### Overall odds ratio and onset time trajectory of identified complications

To assess the overall OR of identified complications, we calculated the 10-year OR for each disease using logistic regression on all patient records, adjusting for age, race, smoking, alcohol use and pre-pregnancy health conditions. Additionally, we performed survival analyses to examine the onset trajectories of these complications. We recorded the time from the first PE diagnosis (cases) or first pregnancy diagnosis (controls) to the onset of each complication as the survival time. We plot Kaplan-Meier curves, and calculated log-rank p-values using the R package “survminer.” Patients who did not follow up before the event of interest were censored.

### Assess the effect of racial disparity and PE severity in identified complications

To investigate potential racial disparities in the effects of PE among the UMich data, we performed a comparative analysis between Caucasians (71.3%) and African Americans (13.8%), excluding other races due to their low representation (<15% combined). We calculated PE ORs on each identified complication, and plotted KM curves for cases and controls in each racial group, as done for all patients.

We also examined whether PE severity influences the risk of complications. Patients were categorized into severe and mild PE groups based on ICD codes: the severe group included those with any severe PE diagnosis during pregnancy, while the mild group had none. We then computed the OR for complications and plotted survival curves for both groups.

### External validation of significant complications using UKBiobank and Cedar Sinai data

To validate the identified complications after PE and the effect of racial disparity and PE severity, we calculated the OR of these complications in *UK Biobank* and Cedar Sinai database, following a similar process. A binary variable indicating the presence of complication after the patient’s first PE (for cases) or normal pregnancy (for controls) diagnosis serves as the response variable, while PE history, pre-existing complications, social history (smoking and alcohol-use status), and race serve as the regressors, the same way as in UMich cohort. To ensure the results are reliable, we assessed the effective size of each complication and removed complications with fewer positive patients to reach 80% power in odds ratio calculation.

### Analytical tools

All analyses were conducted in R programming language v.4.0.3^27^. We used the “touch” package^28^ to standardize ICD9 and ICD10 codes. We used packages “tidyverse”^29^, “lubridate”^30^for data cleaning and “glmnet”^31^, “survminer”^32^, and “qacReg”^33^ for statistical analysis.

### Data Availability Statement

The EHR data from the University of Michigan Precision Health and Cedar-Sinai limited PHI contains sensitive patient information and cannot be made public. Researchers meeting the criteria for using sensitive data may contact the Research Scientific Facilitators at the University of Michigan Precision Health by emailing or visiting https://research.medicine.umich.edu/our-units/data-office-clinical-translational-research/data-access for more information about requesting data access. For Cedars-Sinai limited PHI data, interested researchers may contact inquiries via. UK biobank clinical data are available through the application approval on https://www.ukbiobank.ac.uk/

## Results

### Study overview and patient characteristics

Using three large medical datasets, we are the first to discover and validate the overlooked post-PE complications following a non-hypothesis-based approach and rigorous confounder adjustment. Utilizing EHR from the *UM Medicine Healthcare System*, we incorporated logistic regression and survival analysis to comprehensively investigate the long-term complications in women after PE. We further measured the effect of race and PE severity on identified complications. All results are externally validated using the *UK Biobank and Cedar-Sinai* EHR data. The overall workflow of the study is shown in **Figure 1**.

**Figure 1.**
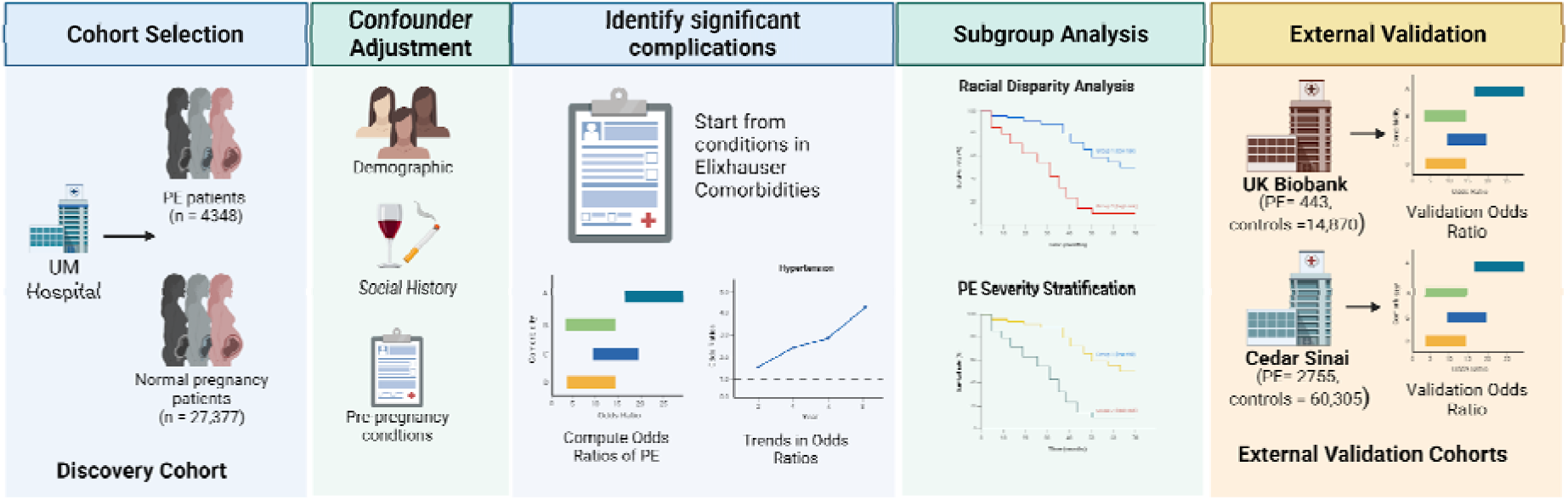
Project workflow. The Discovery Cohort, including PE cases and non-PE controls, was obtained from the *UM Healthcare System*. We select patient demographics, social history and pre-pregnancy conditions as confounders. We then calculate the odds ratio of all Exlihauser comorbidities after PE, with the adjustment of confounders. We plotted the KM curves and calculated the overall OR for identified complications. The results are validated using the UK Biobank and Cedar-Sinai database.

The discovery EHR dataset from *UM Medicine Healthcare System* includes 3,592 preeclampsia cases and 23,040 controls. The validation data from *UK Biobank* include 443 cases and 14,870 controls. The Cedar-Sinai validation data includes 2,755 cases and 60,305 controls. Due to the very limited sample size, we did not use UK Biobank data to validate any stratified results using PE severity or ethnicity. The overall patient characteristics are summarized in **Table 1**. In both discovery and confirmation data, we observed significant differences between the cases and controls in demographic and clinical factors. To account for the differences, we included these factors in the regression models as confounders.

**Table 1:** Patient characteristics in three datasets used in this study.

| Feature |  | UM Data |  |  | Cedar Sinai Data |  |  | UK Biobank Data |  |  |
| --- | --- | --- | --- | --- | --- | --- | --- | --- | --- | --- |
|  |  | Statistics |  | P-value | Statistics |  | p-value | Statistics |  | P-value |
|  |  | Case (n = 3592) | Control (n = 23,040) |  | Case(n= 2755) | Control(n =60305) |  | Case (n = 443) | Control (n = 14,870) |  |
| **Age, Years** |  | 30.647 | 29.694 | <0.001 | 29.290 | 28.309 | <0.001 | 37.630 | 36.290 | <0.001 |
| **Race, Rate** | African American | 603(16.78%) | 2744(11.91%) | <0.001 | 342(12.41%) | 5397(8.95%) | <0.001 | 27(6.1%) | 535(3.6%) | 0.016 |
|  | Caucasian | 2554(71.09%) | 16547(71.82%) |  | 1341(48.68%) | 30333(50.3%) |  | 384(86.7%) | 13383(90%) |  |
|  | Other | 436(12.13%) | 3749(16.27%) |  | 1072(38.91%) | 24574(40.75%) |  | 32(7.2%) | 952(6.4%) |  |
| **Social History, Rate** | Smoking | 1200(33.42%) | 6080(26.39%) | <0.001 | 42(1.52%) | 772(1.28%) | 0.314 | 225(50.8%) | 8030(54%) | 0.219 |
|  | Alcohol | 2225(61.95%) | 13416(58.23%) | <0.001 | 515(18.69%) | 10692(17.73%) | 0.206 | 411(92.7%) | 14082(94.7%) | 0.101 |
| **Pre-existing Disease, Occurrence Rate** | Uncomplicated Diabetes | 198(5.51%) | 309(1.34%) | <0.001 | 123(4.46%) | 597(0.99%) | <0.001 | 3(0.7%) | 24(0.16%) | 0.019 |
|  | Complicated Diabetes | 96(2.67%) | 106(0.46%) | <0.001 | 72(2.61%) | 277(0.46%) | <0.001 | 0(0%) | 3(0.02%) | 1 |
|  | Hypertension Uncomplicated | 420(11.69%) | 484(2.1%) | <0.001 | 168(6.1%) | 271(0.45%) | <0.001 | 3(0.7%) | 24(0.16%) | 0.042 |
|  | Hypothyroidism | 322(8.96%) | 1170(5.08%) | <0.001 | 106(3.85%) | 1079(1.79%) | <0.001 | 3(0.7%) | 34(0.23%) | 0.144 |
|  | Obesity | 588(16.37%) | 1203(5.22%) | <0.001 | 170(6.17%) | 1049(1.74%) | <0.001 | 0(0%) | 12(0.08%) | 1 |
|  | Renal Failure | 55(1.53%) | 99(0.43%) | <0.001 | 18(0.65%) | 30(0.05%) | <0.001 | 0(0%) | 1(0.01%) | 1 |

### Identified six significant complications after PE on the discovery cohort

We first investigated the change in all disease risks each year for 10 years after PE. We adopted the Elixhauser Comorbidity Index and categorized all ICD diagnosis codes into 30 Elixhauser disease categories (see **Method**). For each Elixhauser category, we applied logistic regression with PE as a predictor variable and whether or not diagnosed with the disease during N = 1∼10 years as the response variable, adjusting for other factors such as age, pre-existing conditions, and social history. All features used in this study are listed in **Supplementary Table 3**. If the population of a particular year falls below a reasonable effective size, we exclude that year from the plot to avoid bias (see **Method**). We then calculated the ORs of PE for each Elixhauser category and their confidence intervals (Cis) from these logistic regression models (**Figure 2**). Six complications exhibit statistical significance in more than 4 consecutive years after the first PE (for cases) or normal pregnancy (for controls) diagnosis, during the 10 years. These include uncomplicated hypertension, renal failure, complicated diabetes, uncomplicated diabetes, obesity and hypothyroidism. Notably, the ORs of hypothyroidism after PE are significantly lower than 1 in the first 5 years after patients’ first PE (for cases) or normal pregnancy (for controls) diagnosis, suggesting a potential protective effect of PE on hypothyroidism. However, the ORs show an overall elevating trend as the years go by post-pregnancy to approach 1.

**Figure 2.**
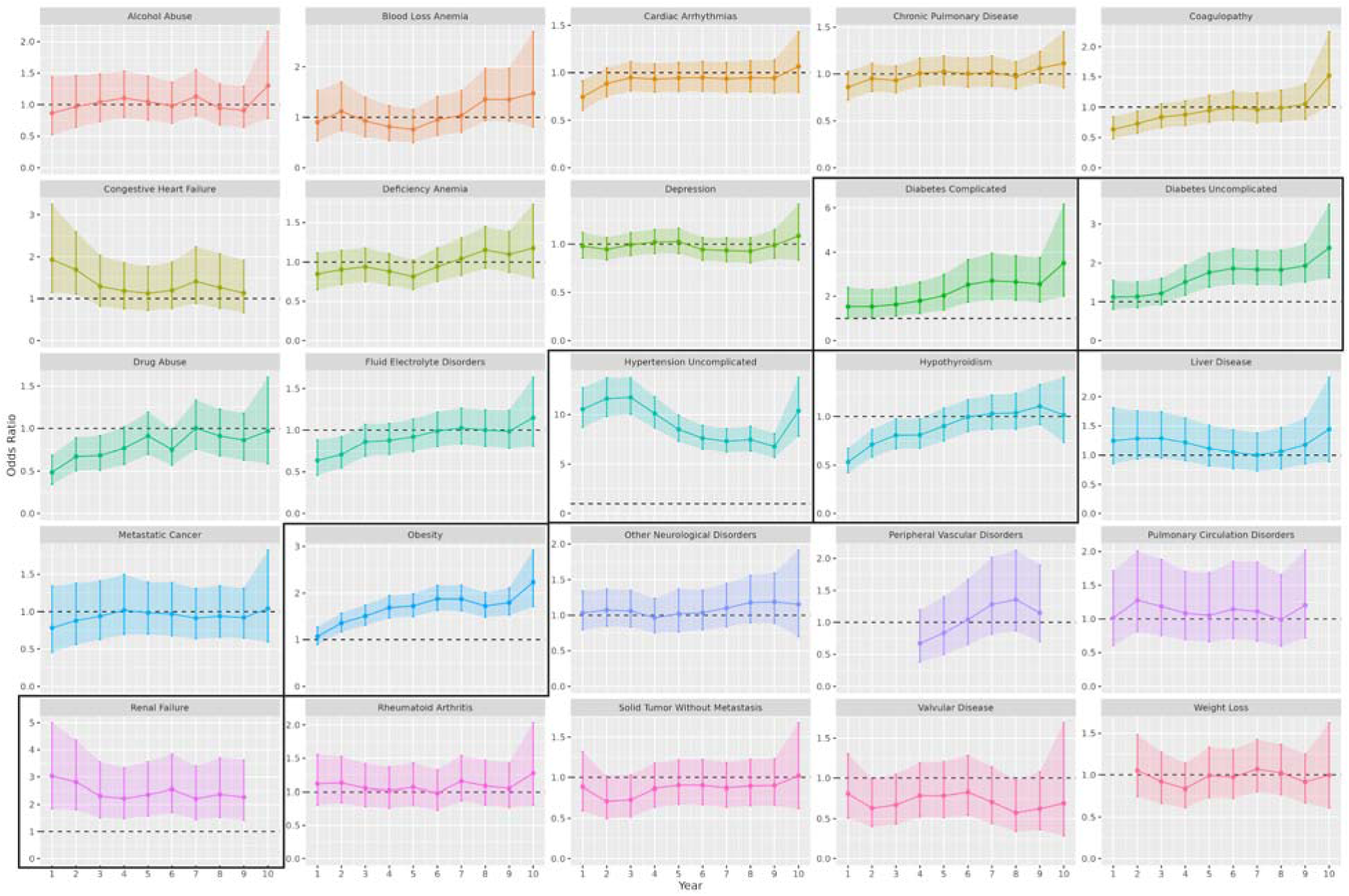
OR trends of all complications defined by Elixhauser Comorbidities. This figure shows the changes in OR of different complications within 1∼10 years after a patient’s first PE (for cases) or normal pregnancy (for controls) diagnosis. For some years certain complications have inadequate effective size for analysis, leading to missing points in the plot. The dashed lines are OR = 1, where ORs above the line indicate higher risks of complications due to PE, and ORs below the line indicate lower risks of complications due to PE. Confidence intervals of the OR are indicated by the colored shadow areas. Plots of complications that are significant for at least 4 consecutive years are boxed.

### Confirmation of identified complications using KM plot and UK Biobank and Cedar-Sinai EMR data

To confirm the identified complications show consistent differences between the PE and controls, we next plotted the Kaplan-Meier (KM) curves and conducted the log-rank test of all six significant complications using UMich data (**Figure 3A**). All selected complications show significant differences between the survival curves of the cases and controls (Log-Rank test, p < 0.05). The difference in hypothyroid occurrence between cases and controls is smaller than other diseases, yet the trend is still significant.

**Figure 3.**
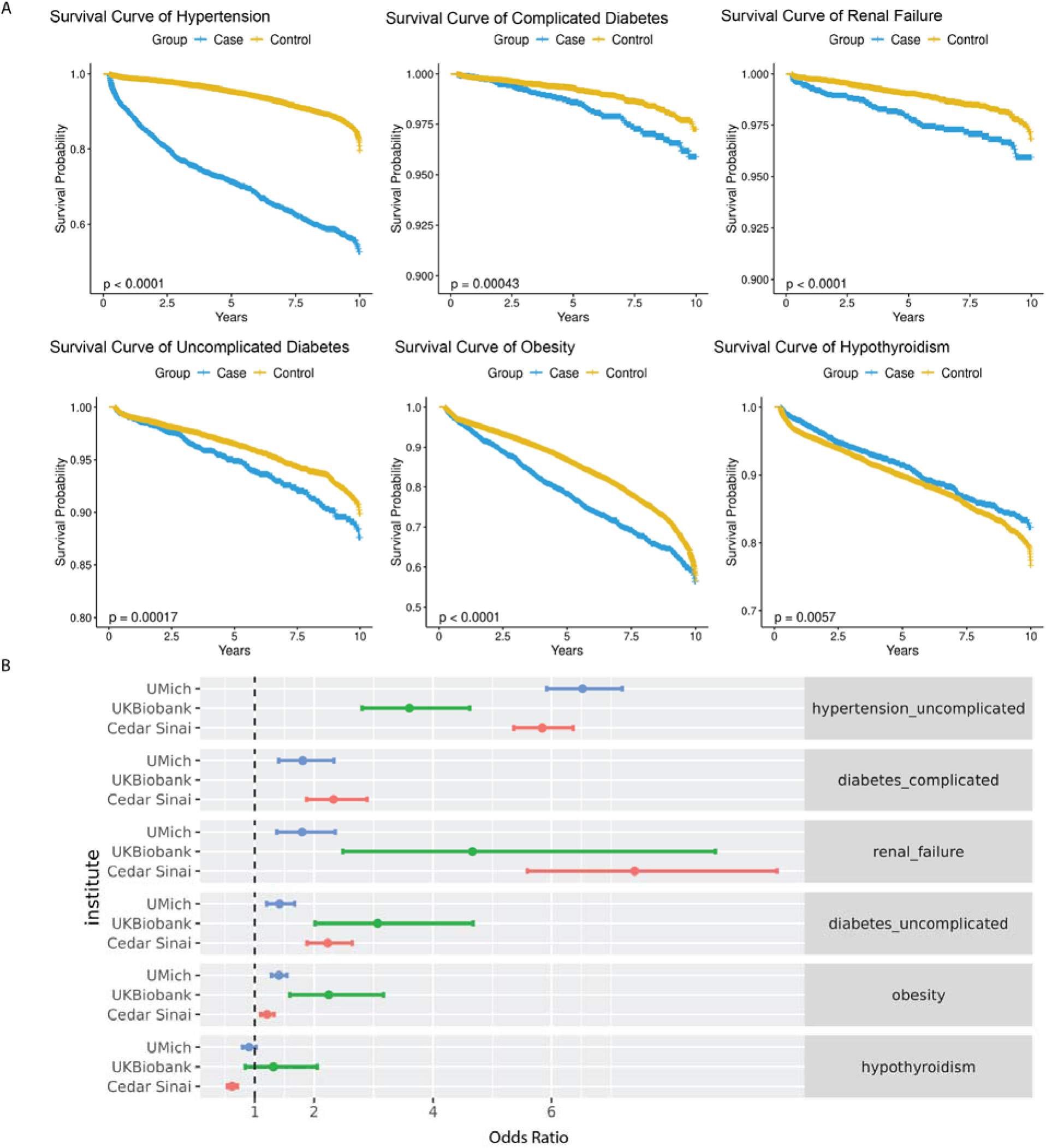
Characterization of significant complications after PE diagnosis. (A) Kaplan-Meier curves of the onset time of significant complications of PE in UMich data. The p-values are calculated from log-rank tests comparing the survival time of cases vs. controls. (B) Odds Ratio(OR) of significant complications after PE across UMich, UK Biobank and Cedar Sinai datasets, adjusted for confounders(patient age, race, smoking, alcohol usage and pre-pregnancy conditions).

To validate the significance of identified complications, we calculated their ORs over 10 years in two external datasets: UK Biobank and Cedar-Sinai data and compared them with those from UMich data in **Figure 3B (Supplementary Table 4)**. For the two validation datasets, we removed diseases with not enough positive cases to reach 80% of power and adjusted for confounders the same way as in UMich data from the figure (see **Methods**). Most ORs for the complications have consistent and positive trends across the three datasets, except for hypothyroidism. Uncomplicated hypertension is the most consistent complication after PE, with OR above 5 in UMich and Cedar-Sinai data and above 3 in UK Biobank. Renal failure, while having positive ORs in the three cohorts, does vary quite a bit in terms of average ORs between cohorts. Uncomplicated diabetes, complicated diabetes, and obesity all show consistently positive ORs in the three datasets, although the OR values are lower than those from hypertension and renal failure.

Hypothyroidism shows negative ORs in both UMich (OR = 0.903, CI: 0.797-1.023) and Cedar-Sinai data (OR = 0.621, CI: 0.579-0.664), although not significant in UK Biobank data due to the small size of PE patients. This suggests a potential protective effect of PE on this complication, consistent with the survival plot in **Figure 3A**. Interestingly, despite the distinct survival curves, the OR of hypothyroidism is not significant in the UMich cohort (p = 0.109). One explanation is that more control patients develop hypothyroidism shortly after delivery but the difference eventually became insignificant later. This is supported by the year-specific odds ratio plot (**Figure 2**), where OR of hypothyroidism is only significant for four consecutive years and with gradually increasing p-values (p=8.76e-8, p = 7.28e-4, p = 0.026, p= 0.026, respectively).

### Comparing the effects of mild and severe PE on later complications

As a heterogeneous disease, PE has various clinical subtypes, among which the severity of PE significantly affects the disease prognosis, outcomes and management^34^. We next explored how severe PE and mild PE may impact patients’ complications differently in the long term. We plotted the KM curves of severe PE patients, mild PE patients and controls for each complication (**Figure 4A**). Several complications show up significantly earlier in severe PE patients than in mild PE patients in several complications, including hypertension, renal failure, uncomplicated diabetes and obesity. This trend is most pronounced for hypertension. Interestingly, for hypothyroidism both severe and mild PE patients show lower rates of occurrences post-pregnancy (p=0.012), particularly for mild PE.

**Figure 4:**
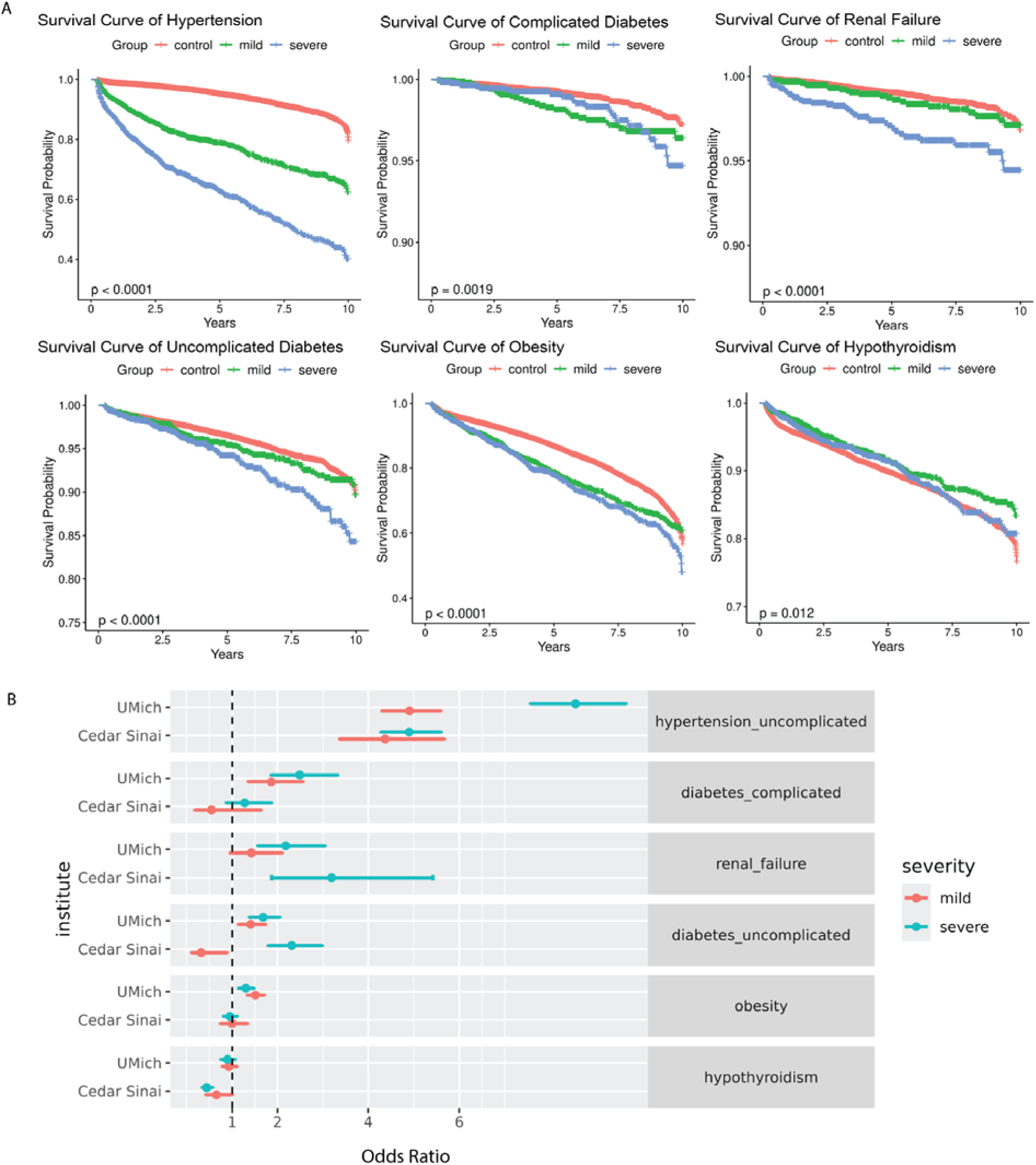
Different effects of severe and mild PE on identified complications. (A) Onset time of complications within 10 years after severe PE, mild PE and normal pregnancies(controls) from the UMich dataset. The p-values were calculated from log-rank tests. (B) The odds ratio and 95% confidence intervals of each complication after PE by severity in UMich and Cedar Sinai datasets

We also looked into the 10-year overall OR of these complications in the UMich cohort and the Cedar-Sinai external datasets within 10 years after PE (**Figure 4B**). We eliminated UK Biobank in this analysis due to the small number of patients with mild vs severe PE records. The trend of changes for ORs of complications between severe vs mild PEs is overall consistent with KM curves. This is particularly the case for hypertension, complicated and uncomplicated diabetes and renal failure, where patients with severe PE have much higher OR compared to those with mild PE. However, the averaged ORs aren’t obviously different in obesity and hypothyroidism OR between severe and mild PE, due to the reduced granularity in this form of data representation compared to KM curves.

### The racial disparity in later complications among PE patients

Further, we examined the possible racial disparity among the identified complications. We compared Caucasian and African American patients, the two most abundant racial groups in the UMich dataset, accounting for 71.7% and 12.6% populations respectively. We performed similar analyses on the two racial groups and plotted their KM curves (**Figure 5A**), showing the onset time of complications in both case and control groups by race. Caucasian patients have lower risks of most complications including hypertension, uncomplicated and complicated diabetes and obesity. However, the differences between cases and controls are more pronounced in Caucasians than African Americans, except for renal failure and hypothyroidism. Noticeably, while in African American patients, PE significantly protects the onset of hypothyroidism compared to controls (Log-Rank test, p = 0.0012), in Caucasians PE does not show a significant effect (p = 0.61).

**Figure 5.**
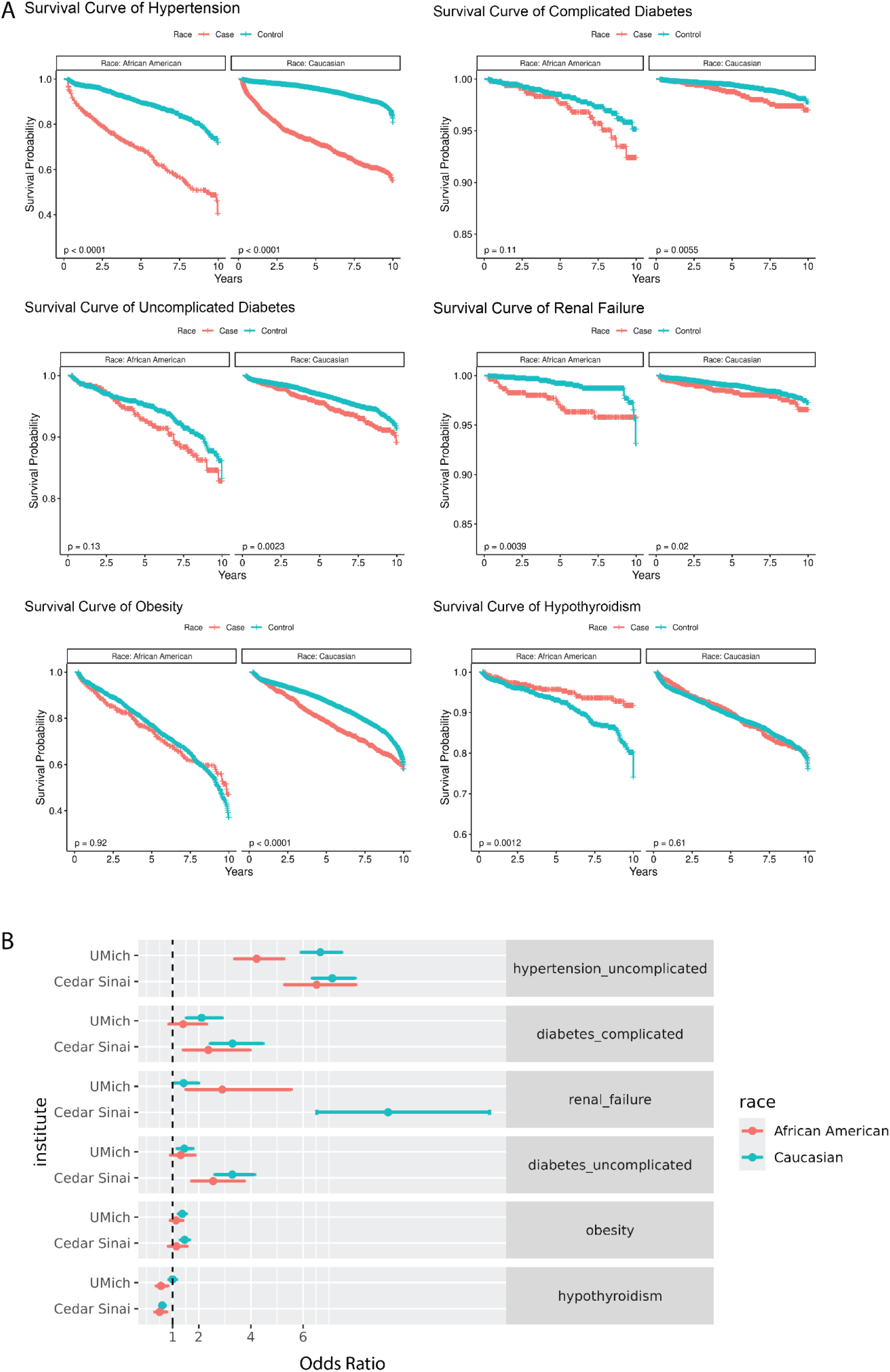
Racial disparities exhibited by complications after PE. (A) Kaplan-Meier(KM) curves of the onset time of complications w.r.t. African Americans and Caucasians in UMich data, separated by PE cases and controls. The p-values were calculated from log-rank tests comparing the survival time of Caucasians and African Americans in cases and controls, respectively. (B) Odds ratios of complications due to PE w.r.t. African Americans and Caucasians in UMich and Cedar Sinai datasets. Complications lacking enough cases to achieve 80% statistical power were excluded from the analysis.

We next investigated the ORs of these complications up to 10 years after PE, in the UMich cohort and the Cedar-Sinai external dataset (**Figure 5B**). We eliminated UK Biobank in this analysis due to the very small percentage of African American patients. For Caucasian patients, most complications remain significant in both datasets, except hypothyroidism results from UMich data. For African Americans, however, PE does not appear to affect the OR of obesity in both datasets (**Figure 5B**). For subsequent complications including hypertension, obesity, complicated and uncomplicated diabetes after PE exposure, their ORs of are all smaller in African Americans than Caucasians. For the details, hypertension has OR = 4.221, CI = [3.387, 5.260] in African Americans and mean OR = 6.662, CI = [5.927, 7.071] in Caucasians.

Obesity has a mean OR = 1.126, CI = [0.916, 1.383] in African Americans, and mean OR = 1.367, CI = [1.226, 1.446] in Caucasians. Complicated diabetes has OR = 1.401, CI = [0.859, 2.285] in African American patients and OR = 2.106, CI = [1.524, 2.484] in Caucasians. For uncomplicated diabetes, OR=1.401, CI=[0.859, 2.285]) in African American patients but OR = 2.106, CI = [1.524, 2.484] in Caucasians. Interestingly, the potential protective effect of PE against hypothyroidism is significant in African Americans (OR = 0.554, CI = [0.375, 0.819]) but not Caucasians(OR = 0.996, CI = [0.865, 1.070]), consistent with the results in **Figure 5A**.

## Discussion

This study is the first to employ a non-hypothesis-based approach and rigorous statistical analysis to comprehensively investigate and validate the overlooked long-term effects of PE using three large medical datasets. We successfully identified and validated 6 complications affected by PE: uncomplicated hypertension, uncomplicated diabetes, complicated diabetes, renal failure, obesity, and hypothyroidism. We also investigate how PE severity and race can affect the effect of PE on these long-term complications differently. We summarized the risks of these complications by PE in **Table 2**. These findings can encourage better management and interventions that benefit post-PE patient care.

**Table 2:** Summarization of PE’s effect on later comorbidities.

| Diseases | All | By Severity |  | By Race |  |
| --- | --- | --- | --- | --- | --- |
|  | All PE cases | Severe PE cases | mild PE cases | PE in African American patients | PE in Caucasian patients |
| Hypertension | ↑ ↑ | ↑ ↑ | ↑ ↑ | ↑ ↑ | ↑ ↑ |
| Complicated Diabetes | ↑ ↑ | ↑ ↑ | ↑ ↑ | ↑ | ↑ ↑ |
| Renal Failure | ↑ ↑ | ↑ ↑ | - | ↑ | ↑ ↑ |
| Uncomplicated Diabetes | ↑ ↑ | ↑ ↑ | ↑ ↑ | ↑ | ↑ ↑ |
| Obesity | ↑ ↑ | ↑ | ↑ | - | ↑ ↑ |
| Hypothyroidism | ↓ | ↓ | ↓ | ↓ ↓ | ↓ |

↑ ↑ Significantly increased risk in all datasets
↑ Significantly increased risk in one or more datasets
- ↓ ↓ Significantly reduced risk in all datasets - ↓ Significantly reduced risk in one or more datasets - no significant change

The significantly increased risks of uncomplicated hypertension, obesity, uncomplicated and complicated diabetes, and renal failure after PE are consistent in all three datasets(**Figure 3A, 3B**). These results align with previous studies about hypertension^7–9,12,14^, diabetes^7,16,35^, renal failure^7,11,15^, and obesity^7,16,35^ after PE, confirming the reliability of our study. One novel finding of our study is the discovery of the potential protective effect of PE against hypothyroidism shortly after PE. A lower rate of hypothyroidism within the first 4 years after PE is observed in both the OR analysis and KM survival curves, although the rate became insignificant in a longer time frame. As expected, further stratification of PE by severe vs. mild symptoms showed that severe PE has a stronger negative effect on many long-term health conditions than mild PE, except obesity and hypothyroidism.

While the racial disparities of hypertension, diabetes, and obesity have been widely recognized^36–38^, there is a lack of research on disparities in PE’s effect on these subsequent conditions. Here we show that PE may have stronger negative effects on Caucasian patients compared to African American patients for hypertension, uncomplicated and complicated diabetes and obesity, despite that the African American population has higher rates of these conditions in both PE-exposed and control groups. This potentially indicates that Caucasians are more sensitive to PE’s inducing effect on these subsequent conditions, compared to African Americans.

Strikingly, the protective effect of hypothyroidism is much more pronounced in African American patients than Caucasians (**Figure 4A**). One previous study identified an increased risk of subclinical hypothyroidism in PE patients^39^, yet one year later a bigger study disagreed and reported no association between PE and subclinical hypothyroidism^40^. In addition, no previous study has been done to investigate the effect of PE on subsequent hypothyroidism in African American patients. One review study suggested that Caucasian patients were generally referred for consultation for hypothyroidism at a younger age compared to African American patients and that non-Caucasian patients have a higher probability of being underdiagnosed^41^. Therefore, it remains to be confirmed whether the racial disparities in hypothyroidism after PE are primarily due to genetic differences or bias in the timing of diagnosis between the two races.

Our study has several notable strengths compared to previous studies on the long-term effects of PE. Methodologically, we utilized a non-hypothesis-based approach to identify any association between complications and PE, allowing for a comprehensive investigation beyond hypothesis-driven approaches to allow the discovery of overlooked associations. We adjusted the analysis for pre-PE medical histories, minimizing the bias due to pre-existing conditions as many of them are indeed associated with PE. This addressed the pitfall in traditional case-control matching studies with only t-tests as statistical evidence^10,13,14,42,43^. In each step of our analysis, we conducted rigorous inference that ensured sufficient sample size and statistical power. We adopted a discovery-confirmation research strategy from two different datasets, strengthening our findings’ generalizability and reliability. There are few previous studies on the racial disparities of PE’s long-term adverse effects. Our study fills the research gap in this topic, revealing the association between race and PE’s long-term effects. Overall, we extended the PE research to a novel dimension, encouraging future work from a racial perspective and promoting precision aftercare of PE, which is important given the known impact of systemic racism on maternal health outcomes^44^.

While our study has provided valuable insights into the long-term effects of PE, it is important to acknowledge the limitations. A key issue is accuracy in cohort selection, as PE diagnosis criteria have changed over time and vary by country. ICD coding practices also differ across regions, which might affect data consistency. However, a recent study found that PE diagnosis using ICD-10 codes had an 85.7% positive predictive value, suggesting reasonable consistency^45^.

External validation from the Cedar-Sinai further supports this, though future studies could benefit from manual validation of PE diagnoses with specialists. Furthermore, as a complex disease, the heterogeneity in pathology, symptoms, and trajectory of PE could also add noise to the cohort^46–48^. Lastly, our findings are purely based on EHR data, which has internal limitations. For example, UK Biobank data do not have sufficient African American patients, nor stratification on some categories. Other types of data that perturb the molecular and pathological processes, such as genetics and genomics data^49^, would be beneficial to deepen the mechanistic understanding of PE’s long-term effects.

## Abbreviations

PE: preeclampsia
HER: electronic health record
UMich: University of Michigan
IRB: Institutional Review Board
ICD: International Classification of Diseases
OR: odds ratio
CI: confidence interval
VIF: Variance Inflation Factor

## Authors’ contributions

LXG conceived the project and supervised the study. HZ and XY conducted the UMich data collection, analyzed the data, and wrote the draft. LT revised the code to validate the results.

WX and RL helped with confirmation using UK Biobank data. JC and ZW validate the results using Cedar-Sinai dataset. EL provided clinical assistance. All authors have read and revised the manuscript.

## Acknowledgments

XY is supported by T32GM141746 fellowship by NIGMS. LXG is supported by grants by NIH/NIGMS, R01 LM012373 and R01 LM012907 awarded by NLM, and R01 HD084633 awarded by NICHD. WX and RL are supported by the Neurology Collaborative Pilot Funding Awards awarded by Cedars-Sinai Medical Center.

## Code Availability

The code used in analysis is available at https://github.com/lanagarmire/PE_long_term_effects

