## Supplementary materials for "Discover overlooked complications after preeclampsia from three real-world medical record datasets of over 100,000 pregnancies"

**Supplementary Table 1. Case-control Selection Criteria Based on ICD-10 Codes.**

| Group | Operation | Condition | ICD-10 Codes | ICD-9 Codes |
|---|---|---|---|---|
| **Case** | Inclusion | Preeclampsia | O14 | 642.4, 642.5, 642.7 |
| **Control** | Inclusion | Normal Pregnancy | Z34, Z35, Z36, Z37 | V22 |
| | Exclusion | Pre-eclampsia | O14 | 642.4, 642.5, 642.7 |
| | | Pre-existing hypertension complicating pregnancy | O10 | 642.0, 642.1, 642.2 |
| | | Pre-existing hypertension with pre-eclampsia | O11 | 642.7 |
| | | Gestational edema and proteinuria without hypertension | O12 | 646.1, 646.2 |
| | | Gestational hypertension without significant proteinuria | O13 | 642.3 |
| | | Eclampsia | O15 | 642.6 |
| | | Unspecified maternal hypertension | O16 | 642.3, 642.9 |

Note: The listed ICD code hierarchically includes all ICD codes starting with it (e.g. O14 includes O14.1, O14.12, O14.3, etc.).

**Supplementary Table 2. ICD-10 Codes for Elixhauser Comorbidities.**

| Comorbidity | ICD-10 Codes |
|---|---|
| **Congestive heart failure** | I09.9, I11.0, I13.0, I13.2, I25.5, I42.0, I42.5 - I42.9, I43.x, I50.x, P29.0 |
| **Cardiac arrhythmias** | I44.1 - I44.3, I45.6, I45.9, I47.x - I49.x, R00.0, R00.1, R00.8, T82.1, Z45.0, Z95.0 |
| **Valvular disease** | A52.0, I05.x - I08.x, I09.1, I09.8, I34.x - I39.x, Q23.0 - Q23.3, Z95.2 - Z95.4 |
| **Pulmonary circulation disorders** | I26.x, I27.x, I28.0, I28.8, I28.9 |
| **Peripheral vascular disorders** | I70.x, I71.x, I73.1, I73.8, I73.9, I77.1, I79.0, I79.2, K55.1, K55.8, K55.9, Z95.8, Z95.9 |
| **Hypertension, uncomplicated** | I10.x |
| **Hypertension, complicated** | I11.x - I13.x, I15.x |
| **Paralysis** | G04.1, G11.4, G80.1, G80.2, G81.x, G82.x, G83.0 - G83.4, G83.9 |
| **Other neurological disorders** | G10.x - G13.x, G20.x - G22.x, G25.4, G25.5, G31.2, G31.8, G31.9, G32.x, G35.x - G37.x, G40.x, G41.x, G93.1, G93.4, R47.0, R56.x |
| **Chronic pulmonary disease** | I27.8, I27.9, J40.x - J47.x, J60.x - J67.x, J68.4, J70.1, J70.3 |
| **Diabetes, uncomplicated** | E10.0, E10.1, E10.9, E11.0, E11.1, E11.9, E12.0, E12.1, E12.9, E13.0, E13.1, E13.9, E14.0, E14.1, E14.9 |
| **Diabetes, complicated** | E10.2 - E10.8, E11.2 - E11.8, E12.2 - E12.8, E13.2 - E13.8, E14.2 - E14.8 |
| **Hypothyroidism** | E00.x - E03.x, E89.0 |
| **Liver disease** | B18.x, I85.x, I86.4, I98.2, K70.x, K71.1, K71.3 - K71.5, K71.7, K72.x - K74.x, K76.0, K76.2 - K76.9, Z94.4 |
| **Peptic ulcer disease, excluding bleeding** | K25.7, K25.9, K26.7, K26.9, K27.7, K27.9, K28.7, K28.9 |

| | |
|---|---|
| **AIDS/HIV** | B20.x - B22.x, B24.x |
| **Lymphoma** | C81.x - C85.x, C88.x, C96.x, C90.0, C90.2 |
| **Metastatic cancer** | C77.x - C80.x |
| **Solid tumor without metastasis** | C00.x - C26.x, C30.x - C34.x, C37.x - C41.x, C43.x, C45.x - C58.x, C60.x - C76.x, C97.x |
| **Rheumatoid arthritis/collagen vascular diseases** | L94.0, L94.1, L94.3, M05.x, M06.x, M08.x, M12.0, M12.3, M30.x, M31.0 - M31.3, M32.x - M35.x, M45.x, M46.1, M46.8, M46.9 |
| **Coagulopathy** | D65 - D68.x, D69.1, D69.3 - D69.6 |
| **Obesity** | E66.x |
| **Weight loss** | E40.x - E46.x, R63.4, R64 |
| **Fluid and electrolyte disorders** | E22.2, E86.x, E87.x |
| **Blood loss anaemia** | D50.0 |
| **Deficiency anemia** | D50.8, D50.9, D51.x - D53.x |
| **Alcohol abuse** | F10, E52, G62.1, I42.6, K29.2, K70.0, K70.3, K70.9, T51.x, Z50.2, Z71.4, Z72.1 |
| **Drug abuse** | F11.x - F16.x, F18.x, F19.x, Z71.5, Z72.2 |
| **Psychoses** | F20.x, F22.x - F25.x, F28.x, F29.x, F30.2, F31.2, F31.5 |
| **Depression** | F20.4, F31.3 - F31.5, F32.x, F33.x, F34.1, F41.2, F43.2 |

**Supplementary Table 3. Features in each year group.**

| Feature / Year | 1 | 2 | 3 | 4 | 5 | 6 | 7 | 8 | 9 | 10 |
|---|---|---|---|---|---|---|---|---|---|---|
| AgeInYears | TRUE | TRUE | TRUE | TRUE | TRUE | TRUE | TRUE | TRUE | TRUE | TRUE |
| PE | TRUE | TRUE | TRUE | TRUE | TRUE | TRUE | TRUE | TRUE | TRUE | TRUE |
| Gestational Diabetes | TRUE | TRUE | TRUE | TRUE | TRUE | TRUE | TRUE | TRUE | TRUE | TRUE |
| AlcoholAbuse | FALSE | FALSE | TRUE | TRUE | TRUE | TRUE | TRUE | TRUE | TRUE | FALSE |
| BloodLossAnemia | TRUE | TRUE | TRUE | TRUE | TRUE | TRUE | TRUE | TRUE | TRUE | TRUE |
| CardiacArrhythmias | TRUE | TRUE | TRUE | TRUE | TRUE | TRUE | TRUE | TRUE | TRUE | TRUE |
| ChronicPulmonaryDisease | TRUE | TRUE | TRUE | TRUE | TRUE | TRUE | TRUE | TRUE | TRUE | TRUE |
| Coagulopathy | TRUE | TRUE | TRUE | TRUE | TRUE | TRUE | TRUE | TRUE | TRUE | TRUE |

| | | | | | | | | | | |
|---|---|---|---|---|---|---|---|---|---|---|
| CongestiveHeartFailure | TRUE | TRUE | TRUE | TRUE | TRUE | TRUE | TRUE | TRUE | TRUE | TRUE |
| DeficiencyAnemia | TRUE | TRUE | TRUE | TRUE | TRUE | TRUE | TRUE | TRUE | FALSE | FALSE |
| Depression | TRUE | TRUE | TRUE | TRUE | TRUE | TRUE | TRUE | TRUE | TRUE | FALSE |
| DiabetesComplicated | TRUE | TRUE | TRUE | TRUE | TRUE | TRUE | TRUE | TRUE | TRUE | TRUE |
| DiabetesUncomplicated | TRUE | TRUE | TRUE | TRUE | TRUE | TRUE | TRUE | TRUE | TRUE | TRUE |
| DrugAbuse | TRUE | TRUE | TRUE | TRUE | TRUE | TRUE | TRUE | TRUE | TRUE | FALSE |
| FluidElectrolyteDisorders | TRUE | TRUE | TRUE | TRUE | TRUE | TRUE | TRUE | TRUE | TRUE | TRUE |
| HypertensionComplicated | TRUE | TRUE | TRUE | TRUE | TRUE | TRUE | TRUE | TRUE | TRUE | TRUE |
| HypertensionUncomplicated | TRUE | TRUE | TRUE | TRUE | TRUE | TRUE | TRUE | TRUE | TRUE | TRUE |
| Hypothyroidism | TRUE | TRUE | TRUE | TRUE | TRUE | TRUE | TRUE | TRUE | TRUE | TRUE |
| LiverDisease | TRUE | TRUE | TRUE | TRUE | TRUE | TRUE | TRUE | TRUE | TRUE | TRUE |
| Lymphoma | FALSE | FALSE | FALSE | FALSE | FALSE | FALSE | FALSE | FALSE | FALSE | TRUE |
| Obesity | TRUE | TRUE | TRUE | TRUE | TRUE | TRUE | TRUE | TRUE | TRUE | TRUE |
| OtherNeurologicalDisorders | TRUE | TRUE | TRUE | TRUE | TRUE | TRUE | TRUE | TRUE | TRUE | FALSE |
| Paralysis | TRUE | TRUE | TRUE | FALSE | FALSE | FALSE | FALSE | FALSE | FALSE | FALSE |
| PeripheralVascularDisorders | TRUE | TRUE | TRUE | TRUE | TRUE | TRUE | TRUE | TRUE | FALSE | FALSE |
| Psychoses | FALSE | TRUE | TRUE | TRUE | TRUE | TRUE | TRUE | FALSE | FALSE | TRUE |
| PulmonaryCirculationDisorders | TRUE | TRUE | TRUE | TRUE | TRUE | TRUE | TRUE | TRUE | TRUE | TRUE |

| | | | | | | | | | | |
|---|---|---|---|---|---|---|---|---|---|---|
| RenalFailure | TRUE | TRUE | TRUE | TRUE | TRUE | TRUE | TRUE | TRUE | TRUE | TRUE |
| RheumatoidArthritisCollagenVascular | TRUE | TRUE | TRUE | TRUE | TRUE | TRUE | TRUE | TRUE | TRUE | FALSE |
| ValvularDisease | TRUE | TRUE | TRUE | TRUE | TRUE | TRUE | TRUE | TRUE | TRUE | FALSE |
| WeightLoss | TRUE | TRUE | TRUE | TRUE | TRUE | TRUE | TRUE | TRUE | TRUE | TRUE |
| SmokingStatusMapped | TRUE | TRUE | TRUE | TRUE | TRUE | TRUE | TRUE | TRUE | TRUE | TRUE |
| AlcoholUseStatusMapped | TRUE | TRUE | TRUE | FALSE | FALSE | FALSE | FALSE | FALSE | FALSE | FALSE |
| IllegalDrugUserStatusMapped | TRUE | TRUE | TRUE | TRUE | TRUE | TRUE | TRUE | TRUE | TRUE | TRUE |
| SexuallyActiveStatusMapped | TRUE | TRUE | TRUE | TRUE | FALSE | FALSE | FALSE | FALSE | FALSE | FALSE |
| CigarettesYN | TRUE | TRUE | TRUE | TRUE | TRUE | TRUE | TRUE | TRUE | TRUE | TRUE |
| CigarsYN | TRUE | TRUE | TRUE | TRUE | TRUE | TRUE | TRUE | TRUE | TRUE | TRUE |
| IVDrugUserYN | FALSE | FALSE | FALSE | FALSE | FALSE | TRUE | FALSE | FALSE | FALSE | FALSE |
| African American | TRUE | TRUE | TRUE | TRUE | TRUE | TRUE | TRUE | TRUE | TRUE | TRUE |
| Other Races | TRUE | TRUE | TRUE | TRUE | TRUE | TRUE | TRUE | TRUE | TRUE | TRUE |
| Caucasian | TRUE | TRUE | TRUE | TRUE | TRUE | TRUE | TRUE | TRUE | TRUE | TRUE |

**Supplementary Table 4: Odds Ratio and p-values for identified complications after PE in 3 institutions**

| Odds Ratio and p-values for identified complications after PE in 3 institutions | | | | | | |
|---|---|---|---|---|---|---|
| | University of Michigan | | | | | |
| Complications | LogitCoef | CoefStd | P-value | Odds Ratio | CI(low) | CI(high) |
| Uncomplicated Hypertension | 1.875 | 0.05 | 0.00E+00 | 6.522 | 5.918 | 7.188 |
| Uncomplicated diabetes | 0.593 | 0.13 | 4.83E-06 | 1.809 | 1.403 | 2.332 |
| Complicated Diabetes | 0.349 | 0.084 | 3.13E-05 | 1.418 | 1.203 | 1.671 |
| Renal failure | 0.586 | 0.138 | 2.12E-05 | 1.798 | 1.372 | 2.356 |
| Obesity | 0.341 | 0.047 | 2.81E-13 | 1.407 | 1.283 | 1.541 |
| Hypothyroidism | -0.102 | 0.064 | 1.09E-01 | 0.903 | 0.797 | 1.023 |
| | Cedar Sinai | | | | | |
| | LogitCoef | CoefStd | P-value | Odds Ratio | CI(low) | CI(high) |
| Uncomplicated Hypertension | 1.754 | 0.044 | 0.00E+00 | 5.777 | 5.304 | 6.292 |
| Uncomplicated diabetes | 0.801 | 0.086 | 1.52E-20 | 2.228 | 1.882 | 2.638 |
| Complicated Diabetes | 0.844 | 0.111 | 2.29E-14 | 2.326 | 1.873 | 2.890 |
| Renal failure | 2.045 | 0.143 | 1.67E-46 | 7.727 | 5.841 | 10.223 |
| Obesity | 0.184 | 0.047 | 9.60E-05 | 1.203 | 1.096 | 1.319 |
| Hypothyroidism | -0.477 | 0.069 | 5.30E-12 | 0.621 | 0.542 | 0.711 |
| | UKBiobank | | | | | |
| | LogitCoef | CoefStd | P-value | Odds Ratio | CI(low) | CI(high) |
| Uncomplicated Hypertension | 1.281 | 0.127 | 5.36E-24 | 3.601 | 2.809 | 4.617 |
| Uncomplicated diabetes | 1.121 | 0.215 | 1.74E-07 | 3.069 | 2.015 | 4.673 |
| Complicated Diabetes | 2.126 | 0.488 | 1.32E-05 | 8,381 | 3.22 | 21.812 |
| Renal failure | 1.54 | 0.321 | 1.67E-06 | 4.665 | 2.486 | 8,751 |
| Obesity | 0.809 | 0.176 | 4.09E-06 | 2.246 | 1.59 | 3.171 |

| | | | | | | |
|---|---|---|---|---|---|---|
| Hypothyroidism | 0.273 | 0.227 | 2.30E-01 | 1.314 | 0.841 | 2.051 |

**Supplementary 5: Odds ratios and p-values of PE by severity**

| mild PE | | | | | | | | | | | | |
|---|---|---|---|---|---|---|---|---|---|---|---|---|
| | University of Michigan | | | | | | Cedar Sinai | | | | | |
| Complications | LogitCoef | CoefStd | P-value | Odds Ratio | CI Low | CI High | LogitCoef | CoefStd | P-value | Odds Ratio | CI Low | CI High |
| Uncomplicated Hypertension | 1.59 | 0.067 | 5.52E-126 | 4.906 | 4.300 | 5.592 | 1.463 | 0.133 | 4.32E-28 | 4.319 | 3.328 | 5.605 |
| Uncomplicated diabetes | 0.341 | 0.106 | 1.28E-03 | 1.407 | 1.143 | 1.731 | -1.140 | 0.522 | 2.88E-02 | 0.320 | 0.115 | 0.889 |
| Complicated Diabetes | 0.621 | 0.161 | 1.16E-04 | 1.861 | 1.357 | 2.551 | -0.602 | 0.558 | 2.81E-01 | 0.547 | 0.183 | 1.635 |
| Renal failure | 0.352 | 0.199 | 7.68E-02 | 1.422 | 0.963 | 2.100 | -16.3425 | 1243.567 | 0.989515 | 0.000 | 0.000 | NA |
| Obesity | 0.413 | 0.062 | 2.60E-11 | 1.511 | 1.338 | 1.707 | 0 | 0.15 | 9.98E-01 | 1 | 0.745 | 1.342 |
| Hypothyroidism | -0.072 | 0.086 | 4.02E-01 | 0.931 | 0.786 | 1.101 | -0.433 | 0.216 | 4.50E-02 | 0.648 | 0.425 | 0.990 |

| severe PE | | | | | | | | | | | | |
|---|---|---|---|---|---|---|---|---|---|---|---|---|
| | University of Michigan | | | | | | Cedar Sinai | | | | | |
| Complications | LogitCoef | CoefStd | P-value | Odds Ratio | CI Low | CI High | LogitCoef | CoefStd | P-value | Odds Ratio | CI Low | CI High |

| | | | | | | | | | | | | |
|---|---|---|---|---|---|---|---|---|---|---|---|---|
| Uncomplicated Hypertension | 2.146 | 0.062 | 4.55E-260 | 8.552 | 7.572 | 9.655 | 1.574 | 0.068 | 1.69E-117 | 4.826 | 4.224 | 5.514 |
| Uncomplicated diabetes | 0.52 | 0.1 | 2.28E-07 | 1.682 | 1.383 | 2.046 | 0.838 | 0.129 | 7.57E-11 | 2.312 | 1.796 | 2.975 |
| Complicated Diabetes | 0.91 | 0.148 | 6.98E-10 | 2.484 | 1.859 | 3.320 | 0.244 | 0.193 | 2.07E-01 | 1.276 | 0.874 | 1.863 |
| Renal failure | 0.78 | 0.169 | 4.23E-06 | 2.18 | 1.566 | 3.038 | 1.077 | 0.282 | 1.33E-04 | 2.934 | 1.689 | 5.102 |
| Obesity | 0.263 | 0.064 | 4.16E-05 | 1.3 | 1.147 | 1.475 | -0.062 | 0.079 | 4.32E-01 | 0.94 | 0.805 | 1.097 |
| Hypothyroidism | -0.108 | 0.087 | 2.13E-01 | 0.898 | 0.757 | 1.065 | -0.795 | 0.124 | 1.35E-10 | 0.452 | 0.354 | 0.576 |

**Supplementary Table 6: Race-specific Odds Ratio and p-values for identified complications after PE in UM and Cedar Sinai datasets**

| | **Caucasian Patients** | | | | | | | | | | | |
|---|---|---|---|---|---|---|---|---|---|---|---|---|
| | University of Michigan | | | | | | Cedar Sinai | | | | | |
| Complications | LogitCoef | CoefStd | P-value | Odds Ratio | CI(low) | CI(high) | LogitCoef | CoefStd | P-value | Odds Ratio | CI(low) | CI(high) |
| Uncomplicated Hypertension | 1.896 | 0.06 | 7.50E-222 | 6.662 | 5.927 | 7.071 | 1.94 | 0.059 | 6.51E-238 | 6.962 | 6.203 | 7.814 |
| Uncomplicated diabetes | 0.368 | 0.104 | 4.35E-04 | 1.444 | 1.177 | 1.603 | 1.191 | 0.118 | 4.14E-24 | 3.292 | 2.614 | 4.145 |
| Complicated Diabetes | 0.745 | 0.165 | 6.48E-06 | 2.106 | 1.524 | 2.484 | 1.194 | 0.155 | 1.30E-14 | 3.299 | 2.435 | 4.470 |
| Renal failure | 0.351 | 0.172 | 4.17E-02 | 1.421 | 1.013 | 1.688 | 2.201 | 0.18 | 1.98E-34 | 9.036 | 6.351 | 12.856 |
| Obesity | 0.313 | 0.056 | 2.09E-08 | 1.367 | 1.226 | 1.446 | 0.37 | 0.06 | 6.36E-10 | 1.447 | 1.287 | 1.628 |

| | | | | | | | | | | | |
|---|---|---|---|---|---|---|---|---|---|---|---|
| Hypothyroidism | -0.004 | 0.072 | 9.51E-01 | 0.996 | 0.865 | 1.07 | -0.494 | 0.084 | 3.44E-09 | 0.61 | 0.518 | 0.719 |

| African American | | | | | | | | | | | | |
|---|---|---|---|---|---|---|---|---|---|---|---|---|
| | University of Michigan | | | | | | Cedar Sinai | | | | | |
| Complications | LogitCoef | CoefStd | P-value | Odds Ratio | CI(low) | CI(high) | LogitCoef | CoefStd | P-value | Odds Ratio | CI(low) | CI(high) |
| Uncomplicated Hypertension | 1.440 | 0.112 | 0.000 | 4.221 | 3.387 | 5.261 | 1.874 | 0.106 | 1.69E-70 | 6.516 | 5.298 | 8.014 |
| Uncomplicated diabetes | 0.270 | 0.176 | 0.126 | 1.310 | 0.927 | 1.851 | 0.936 | 0.196 | 1.85E-06 | 2.549 | 1.735 | 3.744 |
| Complicated Diabetes | 0.337 | 0.249 | 0.176 | 1.401 | 0.859 | 2.285 | 0.860 | 0.263 | 1.08E-03 | 2.364 | 1.411 | 3.960 |
| Renal failure | 1.065 | 0.331 | 0.001 | 2.900 | 1.517 | 5.545 | 2.187 | 0.324 | 1.40E-11 | 8.908 | 4.724 | 16.798 |
| Obesity | 0.119 | 0.105 | 0.258 | 1.126 | 0.917 | 1.384 | 0.129 | 0.157 | 4.09E-01 | 1.138 | 0.837 | 1.547 |
| Hypothyroidism | -0.590 | 0.199 | 0.003 | 0.554 | 0.375 | 0.819 | -0.691 | 0.219 | 1.61E-03 | 0.501 | 0.326 | 0.770 |
